# Fluorescence Spectroscopy for Real-Time Intraoperative Detection of Middle-Ear Cholesteatoma

**DOI:** 10.1101/2025.05.01.25326244

**Authors:** Joackim Mahdjoub, Olivier Gaiffe, Riham Altaisan, Nikolaos Zirganos, Nicolas Passilly, Bruno Wacogne, Emmanuel Ramasso, Laurent Tavernier

## Abstract

**Objective:** To evaluate the potential of autofluorescence spectroscopy as a real-time intraoperative tool for cholesteatoma detection and differentiation from surrounding non-cholesteatoma middle-ear tissues.

**Study Design:** Prospective ex vivo study.

**Setting:** Besançon University Hospital, France (tertiary care center).

**Methods:** Autofluorescence spectral scans were obtained from middle ear biopsies following 405-nm laser excitation. Histopathologic analysis confirmed tissue classification. Spectral data were analyzed using principal component analysis and classified using a quadratic discriminant analysis model.

**Results:** In a cohort of 23 patients (36 tissue samples, 3,787 fluorescence spectra), the classification model achieved an accuracy of 94.5%, with a sensitivity of 94.7% and a specificity of 94.2%. All biopsies were correctly classified as cholesteatoma or non-cholesteatoma.

**Conclusion:** Autofluorescence spectroscopy demonstrated high diagnostic accuracy for cholesteatoma detection. The simplicity and real-time applicability of the method suggest its potential for intraoperative integration, aiming to enhance surgical precision and reduce recurrence rates. Future in vivo validation will be necessary to assess its feasibility in clinical practice.

## Introduction

Cholesteatoma is a progressive and destructive middle-ear pathology characterized by the accumulation of keratinizing squamous epithelium within the tympanic cavity or mastoid^1^. Although its clinical presentation is often benign—typically involving otorrhea and conductive hearing loss—it can lead to serious complications, including ossicular chain erosion, facial nerve palsy, labyrinthine fistula, and even intracranial infections.

Surgical excision remains the only curative treatment, but achieving complete removal is often challenging. Reported rates of residual or recurrent disease range from 10% to 50%, largely due to the intraoperative difficulty of distinguishing cholesteatoma from surrounding healthy structures^2,3^. Current surgical guidance relies primarily on white-light microscopy or endoscopy, which may not provide sufficient contrast between pathological and non-pathological tissue. Finding a real-time intraoperative tool capable of enhancing tissue discrimination could therefore have a significant impact on surgical outcomes.

Several optical diagnostic techniques, including Raman spectroscopy, autofluorescence imaging, and multispectral imaging, have been investigated for middle-ear tissue classification^4–7^. While some studies have explored the use of exogenous fluorescence agents to enhance cholesteatoma visualization, such approaches are limited by regulatory constraints, potential toxicity, and the time required for tissue uptake^8,9^. Among label-free methods, autofluorescence spectroscopy (AFS) has shown particular promise for tissue characterization in both head and neck oncology and otologic surgery^6,10–14^. Prior studies have demonstrated that 405-nm excitation enables discrimination between keratinized and non-keratinized tissues and allows assessment of the optical redox ratio, a parameter closely associated with cellular metabolic activity and particularly relevant in the context of cholesteatoma^4,15^.

The objective of this study was to assess the diagnostic performance of an AFS-based classification model in distinguishing cholesteatoma from non-cholesteatoma middle-ear tissues. Ultimately, the goal is to evaluate its potential for intraoperative application, enabling real-time, label-free tissue discrimination to support more precise and complete surgical resections.

## Materials and Methods

### Study Design and Subjects

This prospective ex vivo study was conducted between 2021 and 2022. It was deemed to fall outside the scope of the Jardé Law and does not meet the definition of research involving human participants as specified in Article L1122-1 of the French Public Health Code. As such, approval from an ethics committee was not required.

The study was registered by the University Hospital of Besançon (CHU de Besançon). Data collection and processing were carried out by a team from the CHU de Besançon in compliance with the European General Data Protection Regulation (GDPR) and in accordance with the guidelines of the French Data Protection Authority (CNIL). All patient data were fully anonymized prior to analysis.

Middle ear biopsies were collected during routine cholesteatoma surgeries, immediately placed in a saline solution, and analyzed using autofluorescence spectral acquisition before being sent for histopathological confirmation.

### Fluorescence Spectroscopic Measurements

Autofluorescence spectral measurements were acquired using a custom-built spectroscopy system equipped with a 405-nm laser and a fiber-optic probe^16^.

Fluorescence signals were recorded across a 2 mm × 2 mm tissue surface using a 200-μm step size, enabling spatially resolved hyperspectral acquisition.

To ensure spatial correspondence between measurement points and tissue architecture, a position-calibrated imaging system was integrated into the setup (**Figure 1, A**). The acquired images were processed with a segmentation algorithm to remove pixels located outside the tissue area, enhancing the reliability of the analysis (**Figure 1, B**).

**Figure 1.**
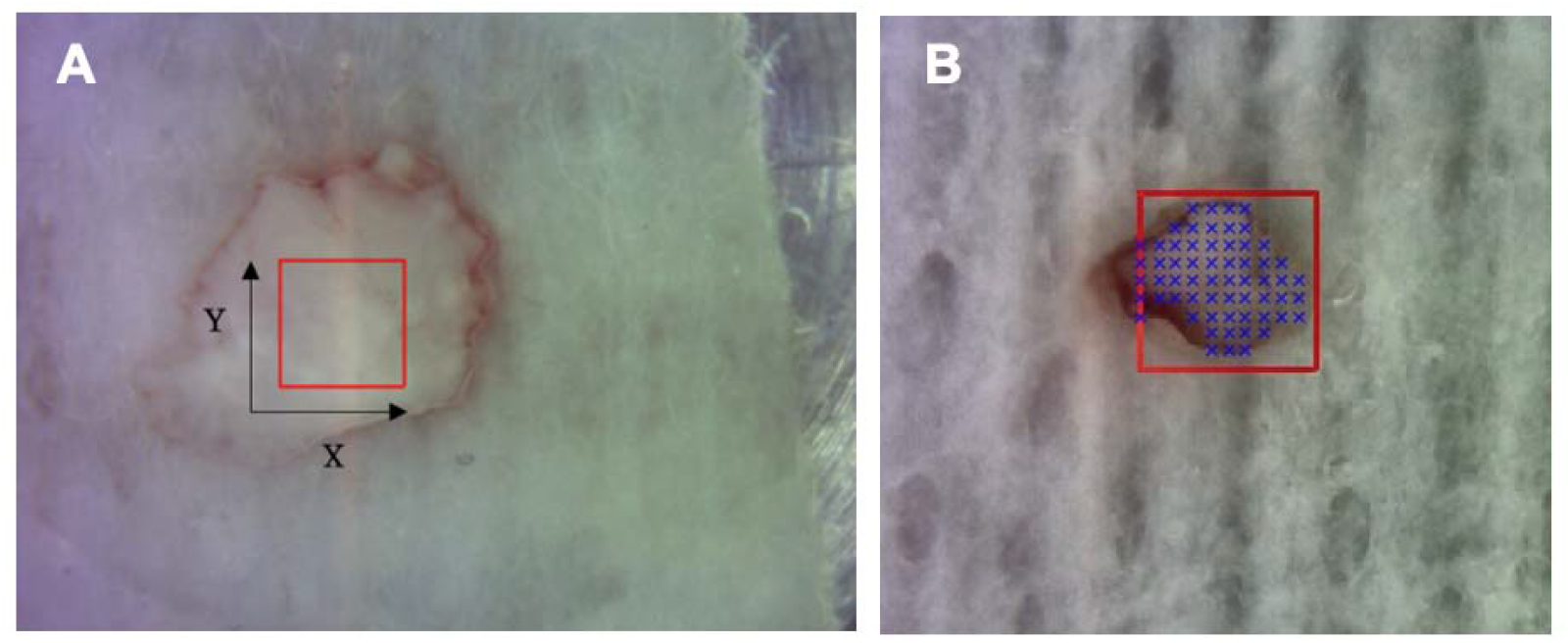
(A) Image of the 4 mm^2^ scan (red square) on a cholesteatoma matrix sample; (B) Segmentation process applied to the calibrated image of a sample, designed to remove data points located outside the sample.

Fluorescence spectra were normalized using the area-under-the-curve (nAUC) to minimize the effects of excitation intensity and reflectivity variation across samples. Spectra exhibiting signal saturation or a signal-to-noise ratio (SNR) < 20 dB were excluded from analysis.

### Data Analysis and Classification

To analyze the high-dimensional spectral data, Principal Component Analysis (PCA) was applied for dimensionality reduction, extracting the most relevant spectral features while minimizing noise and computational complexity. A component selection strategy referred to as the “Select PCs” approach was implemented, which involved the progressive inclusion of principal components only when they improved diagnostic accuracy (Acc). This method enhanced classification performance by improving class separation and reducing the risk of overfitting. The selected components were used as input for a Quadratic Discriminant Analysis (QDA) model to classify each spectrum as either “cholesteatoma” or “non-cholesteatoma middle-ear tissues”.

A 5-fold cross-validation method was implemented to assess model robustness. Final classification of each biopsy was determined by majority voting across its spectra. Diagnostic accuracy, sensitivity, and specificity were calculated, each reported with 95% confidence intervals (CIs). All analyses were performed using *MATLAB R2023a (The MathWorks Inc*., *Natick, MA, USA)*.

## Results

### Population and Sample Characteristics

A total of 23 patients (13 males, 10 females; mean age: 38 years, range: 12–79 years) were included in the study. From these patients, 36 middle-ear tissue samples were collected. Histopathological examination classified the samples into eight distinct categories: cholesteatoma (matrix), cholesteatoma (keratin debris), mucosa, bone, cartilage, connective tissue, muscle, and tympanosclerosis. After preprocessing and quality control, 3,787 fluorescence spectra were retained for analysis. **Table 1** details the distribution of samples and selected spectra across tissue types.

**Table 1.**
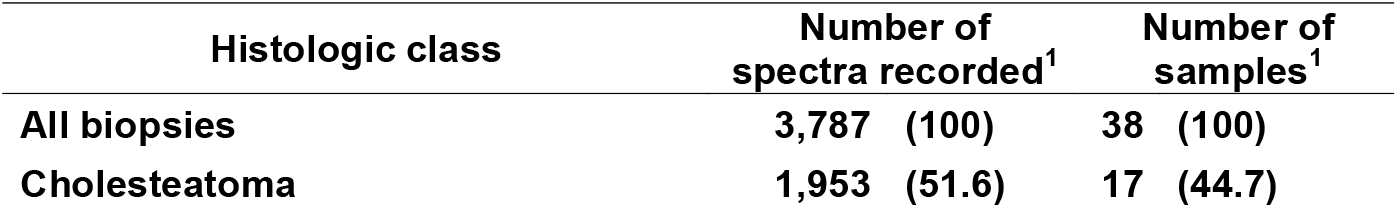
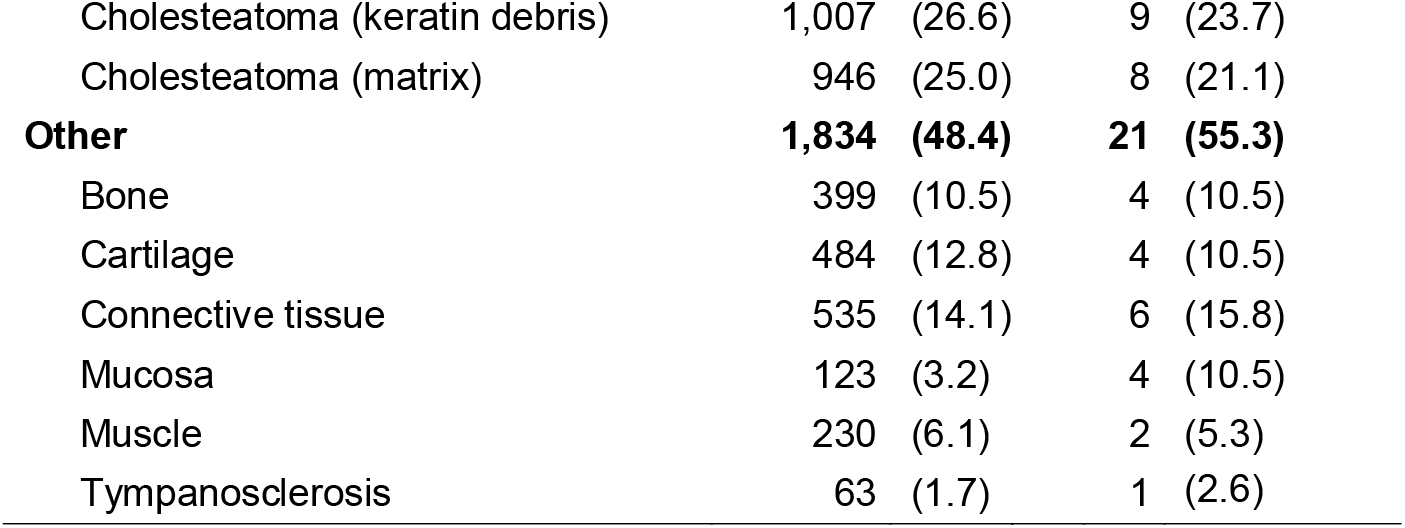
Distribution of selected spectra. ^1^n (%)

### Classification: Cholesteatoma versus Other tissues

The classification model using the “Select PCs” and QDA approach achieved an overall accuracy of 94.5%, with a sensitivity of 94.7% (95% CI: 93.6–95.7%) and a specificity of 94.2% (95% CI: 93.1–95.2%). As illustrated in **Figure 2**, the classification accuracy increased rapidly with the inclusion of the first few principal components, reaching over 93% with just six components. The highest accuracy (94.38%) was observed when using 15 components.

**Figure 2.**
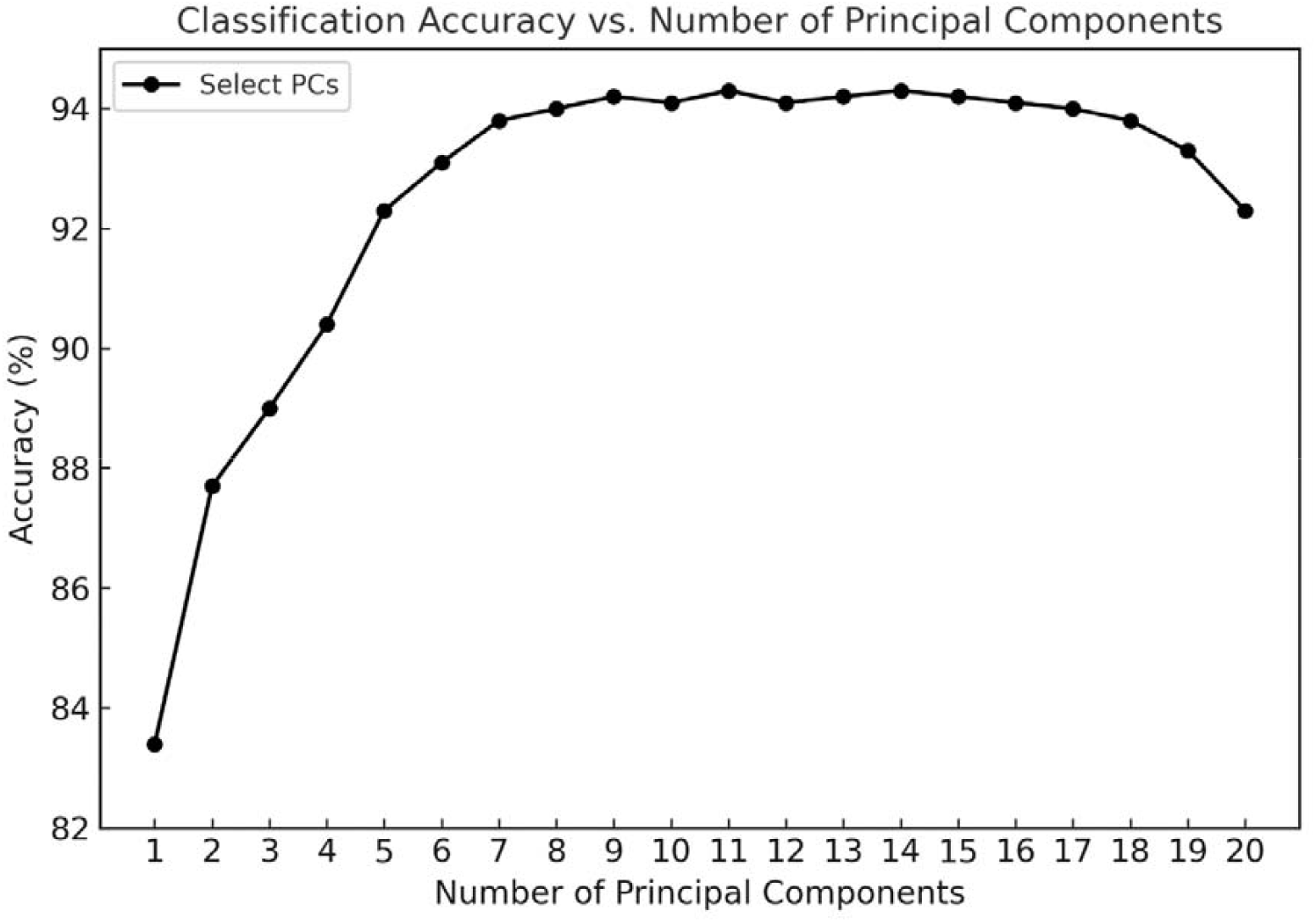
Classification accuracy (%) obtained using a quadratic discriminant analysis (QDA) model as a function of the number of principal components (PCs). The PCs were selected based on their contribution to improving classification performance (Select PCs).

Beyond this point, the performance began to decline slightly or plateau, suggesting that adding more components does not necessarily lead to better classification and may introduce redundancy or noise. Overall, the Select PCs approach enabled the model to achieve high accuracy with fewer components, demonstrating the advantage of targeted component selection over traditional variance-based ordering.

When each biopsy was considered as a whole, all samples were correctly classified as either cholesteatoma or non-cholesteatoma. These results emphasize that selecting PCs based on their contribution to classification performance yields a more accurate and robust model than relying on explained variance alone.

## Discussion

This study presents the first report of a classification technique for middle-ear cholesteatoma based on autofluorescence spectroscopy (AFS) combined with machine learning. Our model achieved a high diagnostic performance, with an accuracy of 94.5%, demonstrating its potential for real-time intraoperative tissue discrimination.

Cholesteatoma surgery is often complicated by the risk of residual disease, mainly due to the difficulty in differentiating pathological tissue from adjacent non-pathological structures during the procedure. In this context, a rapid, reliable, and objective tool to assist in intraoperative identification is critically needed. Our findings support the potential of AFS to address this clinical gap.

Previous optical approaches have explored otoscopic autofluorescence, Raman spectroscopy, and deep learning on otoscopic images. However, most of these studies involved small sample sizes or lacked quantitative validation. For instance, Valdez et al. and Pandey et al.^5,17,18^ evaluated autofluorescence-based tools in pediatric populations and ex vivo tissues, respectively, but included no more than 11 tissue samples and did not report diagnostic performance metrics.

Other groups have focused on using optical image-based classification methods, often involving deep learning. Cavalcanti et al.^19^ trained several classifiers using a smartphone modified for autofluorescence imaging to distinguish normal tympanic membranes from chronic otitis media but obtained only moderate accuracies (50– 79%). Tseng et al.^20^ applied convolutional neural networks (CNNs) to otoscopic images and reported high specificity (96.5%) but only moderate sensitivity (63.6%), which limits their usefulness in reliably detecting disease. Although these otoscopy-based methods may support the early-stage detection of middle ear pathology, they remain unsuitable for the intraoperative identification of residual disease, which requires real-time and high-precision tools.

Few studies have attempted to evaluate intraoperative optical discrimination. In a proof-of-concept study, Wisotzky et al.^7^ combined multispectral imaging with white-light endoscopy in three patients, showing promising but non-quantified results. More recently, Miwa et al.^21^ tested CNNs on intraoperative endoscopic images but reported limited performance, with a sensitivity of 42.3% and specificity of 87.5%.

In contrast, our AFS-based method was designed from the outset with intraoperative application in mind. It combines rapid acquisition, robust spectral processing, and a simple classification model (QDA), enabling potential real-time integration into surgical workflows. This approach represents a significant advancement over previous studies, offering higher performance and clinical relevance.

While the diagnostic results are promising, several limitations must be acknowledged. First, the study was conducted in an ex vivo setting, which does not fully replicate the optical and physiological conditions encountered during surgery, such as tissue perfusion, temperature, and motion. As such, in vivo validation will be necessary to confirm the feasibility and clinical utility of the technique in the operative environment. Second, although 36 tissue samples were analyzed, the number of patients per tissue category remains limited, which may affect the generalizability of the model. Expanding the dataset in future multicenter studies would improve model robustness and allow for the inclusion of additional critical anatomical structures, such as the facial nerve or dura mater, which were not represented here for ethical reasons.

Although we used Quadratic Discriminant Analysis (QDA) for its simplicity and clinical compatibility, future studies could explore more complex classification algorithms, such as support vector machines or deep learning models, which may offer improved performance, especially in more heterogeneous datasets.

In summary, this study demonstrates the feasibility of using autofluorescence spectroscopy combined with machine learning for the differentiation of cholesteatoma from non-cholesteatoma middle-ear tissues. With further validation, this approach could support real-time intraoperative guidance, improve surgical precision, and reduce recurrence rates. These findings provide a strong foundation for future in vivo clinical studies and the development of integrated AFS-based surgical tools.

## Conclusion

This study provides the first evidence to support the use of autofluorescence spectroscopy for cholesteatoma classification, achieving an accuracy of 94.5% on ex vivo samples. Normalized fluorescence spectra enhance the robustness of the system by making it independent of absorption or excitation intensity variations, thus improving the reliability of the method in actual clinical conditions. The real-time applicability and robustness suggest a potential for intraoperative use to improve surgical precision and reduce recurrence rates. Future in vivo validation will be essential to confirm its clinical impact.

## Data Availability

All data produced in the present study are available upon reasonable request to the authors

